# Enhancing Medical Knowledge in Large Language Models via Supervised Continued Pretraining on Clinical Notes

**DOI:** 10.64898/2026.04.02.26350065

**Authors:** Davy Weissenbacher, Moizza Shabbir, Ian M. Campbell, Carl T. Berdahl, Graciela Gonzalez-Hernandez

## Abstract

**Background:** Large language models (LLMs) contain limited professional medical knowledge, as large-scale training on clinical text has not yet been possible due to restricted access.

**Objectives:** To continue pre-training an open-access instruct LLM on de-identified medical notes and evaluate the resulting impact on real-world clinical decision-making tasks and standard benchmarks.

**Methods:** Using 500K de-identified clinical notes from Cedars-Sinai Health System, we fine-tuned a Qwen3-4B Instruct model with supervised learning to generate medical decision-making (MDM) paragraphs from patient presentations, and evaluated it on assigned-diagnosis prediction, in-hospital cardiac-arrest mention detection, and a suite of general and biomedical benchmarks.

**Results:** The fine-tuned model produced MDMs that closely resembled those written by physicians and outperformed the base-instruct model and larger clinically untrained models (Qwen3-32B and Llama-3.1-405B Instruct) on assigned-diagnosis prediction, the task most aligned with its training objective. On the task of detecting in-hospital cardiac arrest mentions, the model initially exhibited mild label collapse, but a brief task-specific fine-tuning stage resolved this issue and allowed it to surpass all competitors. The model also demonstrated global general knowledge retention on biomedical and general-domain evaluation benchmarks compared to the baseline.

**Conclusion:** Supervised full fine-tuning on clinical notes allowed the model to incorporate medical knowledge without sacrificing general-domain abilities, and to transfer this knowledge to unseen biomedical tasks without wholesale loss of general-domain abilities, while revealing collapse-related failure modes that motivate more principled strategies for clinical specialization.

## 1. Introduction

Large Language Models (LLMs) have enabled the first natural language processing systems that can produce coherent and contextually appropriate answers to open-ended questions. This marks a major advance in the field and has established LLMs as the *de facto* standard approach in modern Natural Language Processing. Their remarkable performance stems largely from the ability to transfer real-world knowledge acquired during pre-training. In this phase, models are exposed to a vast corpus of publicly available text and are trained to predict masked or next words within sequences. Each incorrect prediction prompts an adjustment to the internal weights of the model, gradually improving its ability to infer the most probable continuations of text. For example, given the sequence “During an episode of pain, an EKG reveals [***]”, a clinically trained LLM might predict “ST depressions” because it has learned, from repeated co-occurrences in medical text, that this finding is commonly associated with chest pain.

Although the precise mechanisms by which neural networks internalize this knowledge are not fully understood, emerging evidence suggests that LLMs learn structured, grounded representations of entities and their relationships in the real world.^1^ This enables them to generalize knowledge and generate plausible answers to previously unseen questions. As model size increases, LLMs exhibit qualitatively new reasoning abilities:^2^ sufficiently large models display human-like chains of thought–step-by-step reasoning processes that improve both interpretability and task performance.^3^

In medicine, the largest LLMs have demonstrated human-level performance on benchmarks such as MedQA, and have received sufficient media attention to spark growing interest among healthcare professionals in deploying these models in clinical settings.^4^ However, these benchmarks consist primarily of narrowly scoped, synthetic questions that fail to capture the ambiguity and inconsistencies inherent in real-world clinical data.^5^ By focusing mainly on medical licensing examinations and diagnostic reasoning, they overlook a wide range of tasks that are central to hospital operations and everyday clinical practice. When evaluated in more diverse and realistic clinical tasks, their performance declines markedly, revealing a substantial gap that must be addressed before such models can be safely and effectively deployed in healthcare settings.^2,6,7^ This gap limits not only diagnostic performance but also the capacity of current models to reason over longitudinal, ambiguous, or incomplete clinical information—scenarios that dominate routine care.

One major contributor to this gap is insufficient exposure to domain-specific data during pre-training. Electronic Health Records (EHRs), the primary source of clinical documentation, capture the full complexity and nuance of real-world health care, including temporal evolution of disease, comorbidity patterns, and diverse documentation styles. However, patient privacy constraints restrict access to these records to the institutions that maintain them, limiting their general use for model development. Consequently, current open-source models, trained primarily on generic public text, struggle to represent the intricacies of authentic clinical concept relationships, highlighting the critical need for EHR-informed clinical language models.^8^ At the same time, there is limited empirical evidence on how directly training LLMs on real-world clinical notes affects not only task-specific performance but also broader model capabilities and safety.

The goal of this study is to develop and systematically evaluate an instruction-tuned large language model trained on de-identified EHR data, designed to bridge the gap between synthetic medical benchmarks and the complexity of real-world clinical reasoning. We provide a schematic overview of the end-to-end training pipeline and evaluation workflow in Figure 1. Our approach emphasizes both clinical relevance and reproducibility by leveraging real-world notes from a large academic health system and by benchmarking the resulting model against publicly available, instruction-tuned open-source models across multiple tasks and domains.

**Fig. 1:**
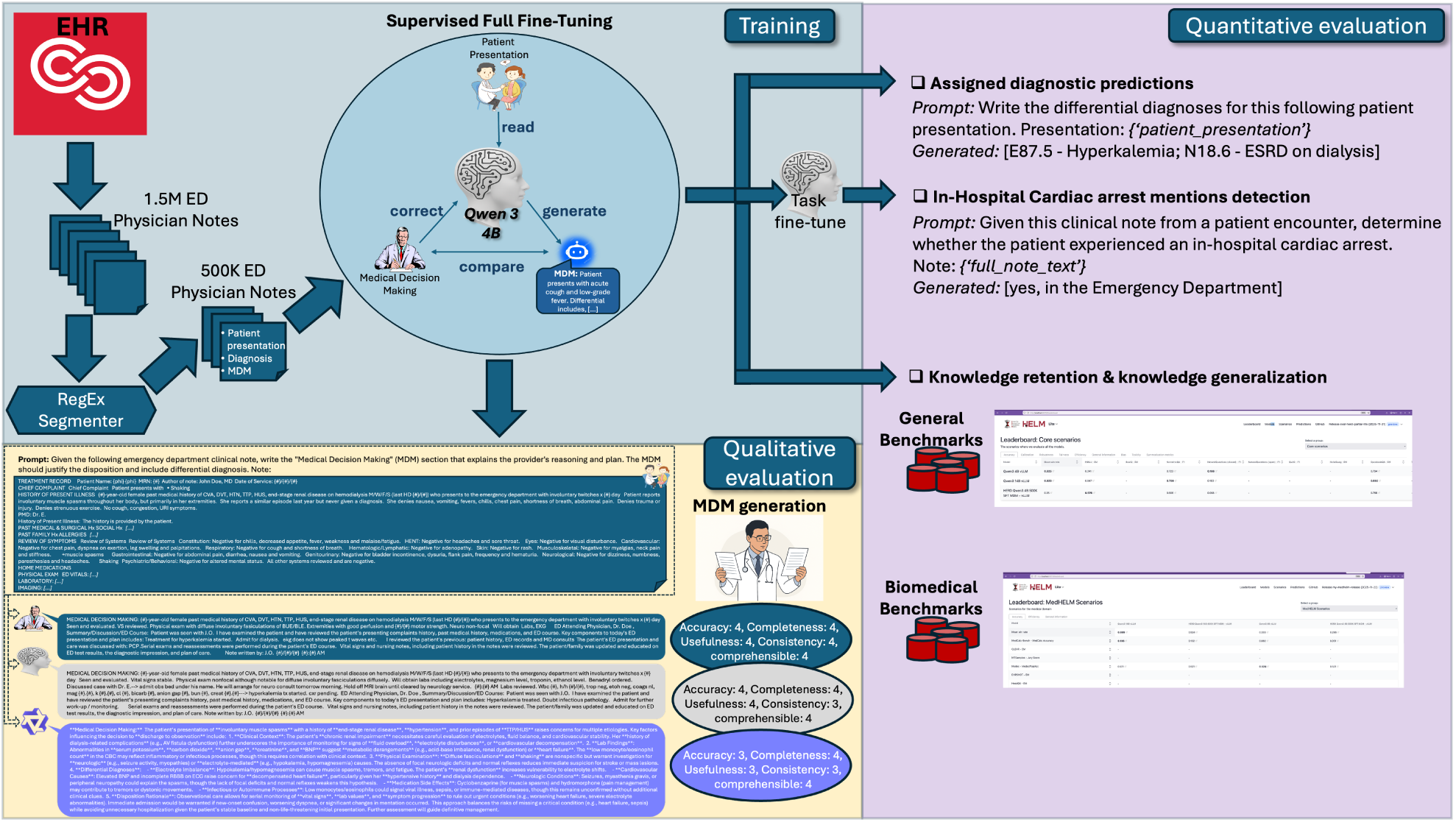
Schematic overview of the end-to-end training pipeline and evaluation workflow.

Among available EHR data sources, this study focuses on Emergency Department (ED) physician notes as a particularly valuable resource for training and evaluating LLMs aimed at enhancing their clinical reasoning and decision support. Emergency physicians manage large numbers of acutely ill patients in an inherently unpredictable and time-constrained environment, where rapid diagnosis and treatment of time-sensitive conditions are essential to reducing morbidity and mortality. During an ED encounter, the attending physician elicits a description of the patient’s symptoms, performs a physical examination, orders and interprets diagnostic tests, and synthesizes findings to formulate a differential diagnosis, with a particular focus on attending or ruling out life-threatening conditions while identifying the most likely underlying causes of the patient’s presentation.^9^ These encounters are documented in ED physician notes, which typically conclude with a *Medical Decision Making (MDM)* or *Assessment and Plan* section that encapsulates the physician’s diagnostic reasoning, clinical judgments, and management plan with supporting evidence.

ED physician notes thus provide a concise, contemporaneous record of clinical reasoning, capturing how physicians integrate subjective symptoms, objective findings, and diagnostic hypotheses under uncertainty to arrive at evidence-based decisions. Because they encompass the full continuum of medical concepts —from presentation and inference to intervention— ED notes constitute an ideal, real-world foundation for studying how LLMs can learn and emulate core processes of clinical reasoning in high-acuity care settings.

To integrate these patterns of medical decision-making into an LLM, we trained a Qwen3-4B instruct model using supervised full fine-tuning on a large corpus of de-identified ED physician notes from our institution. For each encounter, the model was provided with the patient presentation (history, physical examination, and diagnostic testing sections) while withholding the MDM and assigned diagnosis, and was prompted to generate the corresponding MDM paragraph. The training loss was computed by comparing generated and physician-authored MDMs, and all model weights were updated to increase the likelihood of producing MDMs consistent with human-written documentation. Two board-certified emergency physicians (MS and CB) manually evaluated a subset of generated MDMs to confirm that the model had correctly learned the task and to verify basic clinical plausibility.

To assess whether fine-tuning on MDM generation improved broader clinical competence, we evaluated the model against its base instruction-tuned counterpart on two downstream EHR-based tasks: (1) predicting the assigned diagnosis given a patient presentation, which is closely related to MDM generation, and (2) detecting documentation of in-hospital cardiac arrest in randomly selected encounter notes, a more distant task. We further examined biomedical and general-domain performance using the HELM-Lite and MedHELM benchmarks to quantify any overall loss or gain in capability relative to the base model. Taken together, these evaluations provide an empirical assessment of how EHR-based fine-tuning on real-world clinical reasoning signals influences both in-domain and out-of-domain performance, informing the design of future clinically grounded LLMs.

## 2. Related Work

The community has already proposed several approaches to specialize LLMs for the medical domain; however, these efforts remain limited in scope and effectiveness.

Early efforts^10,11^ specialized models by training them from scratch on large collections of real clinical notes from the University of Florida Health Integrated Data Repository combined with general-domain text. While effective, this approach requires substantial computational resources, limiting reproducibility, and lacks reinforcement learning–based alignment, which constrains generalization to unseen tasks and necessitates task-specific fine-tuning. Current studies,^12^ including our work, instead predominately favor continued pretraining to adapt existing foundation models using medical data.

A second key limitation concerns data availability. As early noted by Wornow et al.,^8^ the scarcity of large, open repositories of unstructured EHR text has led the community to rely predominantly on the MIMIC dataset,^13^ resulting in clinical LLMs trained on narrowly scoped datasets with limited note diversity. This landscape remained largely unchanged as of 2025, in their review, Cao et al.^14^ report that a substantial fraction of medical LLMs are trained exclusively on this dataset. In contrast, our work leverages large-scale, real-world EHR data from Cedars-Sinai Medical Center, enabling pretraining our model on 500K clinical notes with the potential to scale to over 5 million patient records. Among existing studies that trained LLMs on real EHR data, Sirrianni et al.^15^ continued pretraining GPT-2 on 374K dental notes using next-token prediction, while Akhondi-Asl et al.,^16^ closer to our study, applied supervised fine-tuning to train a LLaMA-7B model on 34.4K PICU notes for generating differential diagnosis. Although both report improvements over their base models, evaluations are restricted in breadth. In contrast, our study conducts a more comprehensive evaluation, similar to Peng et al.,^11^ by assessing multiple tasks performance as well as potential degradation of the foundation model’s general capabilities.

Finally, an emerging and very active line of work explores multi-modal clinical models, e.g. Jiang et al..^17^ However, we restrict our scope to text-only models and leave multi-modal integration to future work, as it introduces additional algorithmic challenges and significantly increases hardware requirements.

Relative to these prior efforts, our work makes three distinct contributions. First, rather than targeting broad clinical summarization or generic note generation, we focus explicitly on the *Medical Decision Making* section of ED notes as a high-yield representation of real-world clinical reasoning, enabling more direct modeling of diagnostic thought processes under uncertainty. Second, we combine supervised fine-tuning on de-identified EHR text with a systematic, multi-axis evaluation that spans in-domain tasks (MDM generation and diagnosis prediction), distant EHR-based phenotyping (cardiac arrest detection), and established general and biomedical-domain benchmarks, providing a more comprehensive assessment of downstream effects on both clinical and non-clinical capabilities than is typical in existing work. Third, by building on a compact, openly available foundation model (Qwen3-4B instruct) and describing a scalable training and evaluation pipeline on large-scale ED data from a major academic medical center, we offer a practically reproducible framework for developing and vetting clinically grounded LLMs in health systems with similar data resources.

## 3. Materials and Methods

### 3.1. Data collection

This study was conducted using de-identified EHR data and was reviewed by the Cedars-Sinai Medical Center Institutional Review Board, which determined that it met criteria for exemption from human subjects research oversight.

We extracted approximately 5 million emergency department (ED) physician notes from Cedars–Sinai’s electronic health record (EHR) system, representing all notes available at the time of data export. To protect patient privacy, the health system applied a rigorous deidentification process that removed all personally identifiable health information (PHI). Detected PHI spans were replaced with the placeholder token *{phi}*, and all digits—whether part of PHI or not—were substituted with the symbol *{#}*. A major limitation of this work is that only de-identified notes were available for analysis and model development. We programmatically detected and isolated the assigned diagnosis and MDM sections from the patient presentation using regular expressions. Many ED notes adhered to standardized documentation templates, which allowed reliable segmentation with this approach and yielded 511,077 successfully processed notes from the initial corpus. Extending section segmentation to the remaining notes using more advanced approaches, such as prompt-based methods or fine-tuned LLMs, is left for future work.

After isolating the assigned diagnosis section, we restored the corresponding ICD codes removed during de-identification by performing exact string matching between the preferred diagnostic terms in the notes and those in the EHR system’s dictionary table. Since assigned diagnoses often consist of multiple codes, we labeled a note as having an unavailable assigned diagnosis when one or more component diagnoses could not be mapped to ICD-10 or ICD-9 codes. Despite its simplicity, this procedure enabled successful recovery of all ICD codes for the assigned diagnosis in 361,595 of the 511,077 segmented notes. Finally, we randomly partitioned the dataset into 510,327 notes for training, 250 for validation, and 500 for testing. For the MDM generation task, all splits were used as defined above. For diagnostic prediction, only 178 validation notes and 361 test notes had fully recoverable assigned diagnoses and were therefore included in the corresponding experiments.

### 3.2. Training Generative models

Multiple open-source LLMs of varying sizes and performance levels are now available. For our experiments, we selected the Qwen 3 series,^18^ which comprises both dense and mixture of experts models ranging from 0.6 to 235 billion parameters and is specifically trained for reasoning tasks. Due to constraints on available computational resources and training time, we conducted our experiments using the Qwen3-4B model. We used the Transformer Reinforcement Learning library^19^ for its integration with Hugging Face Transformers. We selected the Qwen3-4B Instruct model, which underwent additional reinforcement learning–based fine-tuning to enhance its alignment with human intent and strengthen its reasoning capabilities. We further trained this model using supervised full fine-tuning, providing prompts that instructed it to complete an ED note by generating the appropriate MDM section given a patient presentation. The structure of the prompt is shown in Figure 1. After each completion, the generated MDM was compared with the physician-authored MDM to compute the training loss, and the model weights were updated to minimize this loss in subsequent iterations. We used the library’s default token-level cross-entropy loss function for optimization.

We found that fully supervised training of an instruction-tuned model was particularly challenging, given the need to balance parameter choices that mitigate catastrophic forgetting while operating within limited computational resources. Catastrophic forgetting^20^ is a phenomenon in which a trained model, when fine-tuned on a new task, becomes overspecialized to that task and overwrites the parameters that enabled it to perform previous ones, resulting in a loss of its earlier capabilities. To mitigate this effect, we followed standard recommendations^21^ by augmenting our training set with 10% of general-domain examples^∗^, yielding a total of 734,196 training examples. We further reduced the learning rate to 1e-5 and presented each example only once during training. The model was trained on a single computational node equipped with eight A100 GPUs during 172h. To increase training efficiency, we limited the maximum input sequence length to 6,050 tokens by excluding 18,427 (2.5%) training examples that exceeded this length. All remaining hyperparameters are provided in our publicly available code at github.

We augmented our training set with randomly selected examples from several publicly available sources. The first, the Databricks Dolly 15K dataset,^22^ comprises 15,000 prompt–response pairs created in 2023 by over 5,000 Databricks employees. The prompts cover a wide range of tasks, including open/closed-ended question answering, information extraction, summarization, or creative writing. Contributors were explicitly instructed not to use generative models or copy content from the internet, but to compose original responses. This process promotes linguistic diversity and ensures the quality and factual accuracy of the answers. The second dataset, Super-NaturalInstructions,^23^ was developed through a large community effort in the NLP field to create a comprehensive benchmark comprising over 5 million examples across 1,616 tasks, covering 76 task types and 55 languages. The benchmark compiles existing datasets that were reformatted into a standardized instruction-based schema, including a task definition, input, expected answer(s), and, in some cases, examples of undesired or incorrect outputs. The authors followed a rigorous process to ensure that each task was clearly defined and validated at the task level; however, they did not conduct an individual audit of every example within the corpus. The final dataset, LMSYS-Chat-1M,^24^ consists of approximately one million conversations in multiple languages between anonymous users and 25 large language models (LLMs) of varying sizes, released by the authors in 2023. All personally identifiable information was removed, and conversations flagged as unsafe by the OpenAI moderation API were retained in the original release for academic research. We excluded them from our training set. We treated each turn in a conversation as an individual example; i.e. the model was given the user’s most recent question as input and evaluated against the response generated by the original model.

### 3.3. Evaluation

We performed intrinsic and extrinsic evaluations to assess whether the model acquired medical knowledge during training. Intrinsic evaluation focused on the quality of MDM generation, which is difficult to measure automatically—standard metrics such as ROUGE or BLEU capture only limited linguistic and semantic aspects and do not ensure domain accuracy—so we relied on expert human judgment. Extrinsic evaluation included two automatically measurable tasks: a diagnosis prediction task, closely related to MDM generation, and a detection task, which draws on clinical knowledge but differs in nature. We further assessed performance on additional medical and general-domain benchmarks to gauge the broader impact of fine-tuning on reasoning and language capabilities.

#### 3.3.1. Expert evaluation of model generated MDMs

Following established practices for clinician assessment of AI-generated clinical documentation while aiming to minimize the burden on reviewers,^25,26^ we asked two board-certified ED physicians (MS and CB) to evaluate the quality of model-generated MDMs. We randomly sampled 25 ED encounters from the test setand extracted for each case the physician-authored MDM (gold standard), the MDM generated by the base model (Qwen3-4B Instruct), and the MDM generated by the fine-tuned model. For each encounter, the three corresponding MDMs were presented together in a randomized order and labeled only as versions A, B, and C; reviewers were blinded to note source (physician vs. base model vs. fine-tuned model) throughout the evaluation.

Both physicians independently rated all MDMs using a subset of dimensions from the Physician Documentation Quality Instrument, 9-item version (PDQI-9),^26^ adapted to focus on attributes most relevant to clinical decision-making. Specifically, they scored each note on a 5-point Likert scale (1 = poor, 5 = excellent) along the following dimensions:

*•* **Accuracy**: The note contains no incorrect clinical information.
*•* **Completeness**: The note is comprehensive and captures all clinically relevant issues for the patient.
*•* **Usefulness**: The note is pertinent, offering meaningful information and clinical insight.
*•* **Internal consistency**: Later sections neither contradict nor omit information documented earlier; this dimension captures hallucinations and self-contradictions.
*•* **Comprehensibility**: The note is clear and understandable, without ambiguity or sections that are difficult to interpret.

Ratings were collected individually without discussion. For each dimension, we summarized scores by averaging across encounters and raters for each of the three note sources (gold-standard physician MDMs, base-model MDMs, and fine-tuned-model MDMs). This design allowed us to compare perceived clinical quality across sources and to assess whether fine-tuning improved MDM quality relative to the base model and usual-care physician documentation.

#### 3.3.2. Quantitative Evaluation of Knowledge Transfer and Generalization

*Diagnosis Prediction.* To evaluate whether fine-tuning on MDM generation improved the model’s ability to infer diagnostic labels from patient presentations, we formulated a diagnosis prediction task at the note level. For each note in the test and validation sets with a fully recoverable assigned diagnosis (Section 3.1), the input to the model consisted of the patient presentation (history, physical examination, and diagnostic testing sections), while the output to predict was the set of International Classification of Diseases (ICD-10 or ICD-9) codes with its corresponding preferred term recovered from the note. We used guided decoding to enforce a well-structured JSON output, which simplified parsing. For patient presentations longer than the model’s maximum input length of 6050 tokens, we returned an empty list. Because notes could be assigned multiple diagnosis codes, the task was treated as multi-label classification over the set of distinct ICD codes observed in the dataset. In brief, for each note, we prompted the model with an instruction to infer the patient’s diagnosis from the presentation and required it to output a list of diagnosis descriptions. Model predictions were mapped to ICD codes via exact or fuzzy string matching against the EHR diagnosis dictionary used during data preprocessing, using the same procedure described in the Data Collection Section.

We compared our model against three baselines. First, we evaluated it against the base off-the-shelf Qwen3-4B instruct model, which differs from our model only in the absence of task-specific fine-tuning, allowing us to measure the impact of our training. Second, we included the larger Qwen3-32B instruct model, which shares the same pretraining and instruction-tuning approach as the 4B model but can capture more general knowledge due to its higher capacity. Finally, we benchmarked our model against the Llama 3.1 405B instruct model, a very large model widely used in academic studies.

We evaluated model performance using strict and overlapping micro F1-scores. For the strict metric, we counted a prediction as a true positive (TP) when the model produced the same ICD-10 code as the physician or the exact same lower-cased preferred term^†^. We counted a false positive (FP) when the model predicted a code–term pair not assigned by the physician, and a false negative (FN) when the model missed an assigned condition. We then calculated micro-precision (TP/(TP+FP)), micro-recall (TP/(TP+FN)), and their harmonic mean, the micro-strict F1-score. For the overlapping metric, we applied the same formulas but relaxed the matching rule: we counted a prediction as correct if it had the same ICD-10 code or if the predicted and assigned preferred terms overlapped lexically (i.e., one term appeared as a substring of the other after lower-casing).

We report these metrics for our model and all baselines on the shared test set. Because attending physicians typically justify their assigned diagnoses within the MDM section, this diagnosis prediction task closely mirrors the inferential step that MDM generation is intended to capture, providing a direct test of whether the model’s learned decision-making patterns transfer to structured diagnostic labeling.

Because attending physicians typically justify their assigned diagnoses within the MDM section, this diagnosis prediction task closely mirrors the inferential step that MDM generation is intended to capture, providing a direct test of whether the model’s learned decision-making patterns transfer to structured diagnostic labeling.

*In-Hospital Cardiac Arrest Detection.* The objective of this task was to identify clinical notes documenting an in-hospital cardiac arrest event. We utilized an existing corpus described in,^27^ consisting of 500 patient encounters—235 selected based on predefined ICD codes associated with cardiac arrest and 265 randomly sampled from the Cedars–Sinai EHR system (Epic Systems Corporation, Verona, WI). Each clinical encounter corresponded to a single ED visit and included any ED and inpatient notes generated during that encounter. For this study, we retained only ED notes, hospital discharge summaries, and Code Blue event notes, resulting in a total of 3,822 notes. The dataset was partitioned into 300 encounters for training, 50 for development, and 150 for testing. The corpus exhibits a high degree of class imbalance, as in-hospital cardiac arrests are rare, with only 6.9% (264/3,822) of notes referencing such events.

In our previous study,^27^ through an iterative process, we designed a prompt to perform this task efficiently. The prompt began by defining a cardiac arrest as both the presence of pulselessness and a resuscitation treatment, followed by a brief description clarifying what constitutes an in-hospital cardiac arrest.^28^ For each example, the model received a complete note, preceded by its type and timestamp. Using guided decoding, we enforced a structured JSON output consisting of: (1) excerpts from the note supporting the decision, (2) a brief explanation of the reasoning, and (3) the final label—either no cardiac arrest mentioned or the location of the cardiac arrest within the hospital (*started outside hospital and continued in the ED*, *started in the ED*, *started somewhere else in-hospital*). We use this prompt and guided decoding for our model and all baselines in these experiments.

We compared our model against several baseline systems (Table 3). First, we evaluated it against the best-performing system reported by Vo et al.^27^ (line 5), which used Llama 3.1 405B in a few-shot learning setup. Although no longer the state-of-the-art system, we selected this model due to its scale as a large language model and its consistently strong performance. Second (line 1), we followed the same few-shot setup as done first, but replaced Llama with the off-the-shelf Qwen3-4B Instruct model. This second baseline reflects the standard approach while allowing a direct comparison with our model, since both share the same model size. Third (line 3), we fine-tuned the Qwen3-4B Instruct model on the training set using full supervised fine-tuning (SFT); during inference, we did not include few-shot examples in the prompt, since the model had already seen them during training. We expected this model to outperform the few-shot version because it was directly trained on the task. Fourth (line 4), we continued training our MDM-trained model with supervised fine-tuning on the cardiac arrest detection task using the same training set. Comparing this model with the SFT baseline (line 3) is the most informative comparison as it allowed us to measure the knowledge transfer from learning MDM generation to this new task. When evaluating our own model (line 2), we encountered a practical limitation stemming from an early design choice: to accelerate training on the MDM generation task, we reduced the maximum input length, which prevented the inclusion of few-shot examples in the prompt for cardiac arrest detection. Consequently, this model was conditioned only on task instructions, without demonstrations, placing it at a disadvantage relative to the other baselines. This limitation is not inherent to the approach and could be addressed with additional training time.

We evaluated each model’s performance as a binary classification task, assessing its ability to identify whether a clinical note mentioned an in-hospital cardiac arrest, irrespective of its location. Performance was measured using standard evaluation metrics: precision, recall, and F1-score.

Although conceptually related to clinical reasoning, cardiac arrest detection is operationalized here as a distant, phenotyping-style task that differs substantially from generating MDM narratives. As such, performance on this task probes whether the clinical knowledge and reasoning patterns learned during MDM-focused fine-tuning generalize to structurally different, real-world clinical information extraction problems.

*General Understanding — Benchmark Evaluation.* Standardized LLM benchmark suites group diverse tasks—such as question answering, reasoning, or translation—into curated benchmarks with reference outputs and metrics to assess general capabilities.^29^ To evaluate how clinical fine-tuning affects broader model performance, we compared our fine-tuned Qwen3-4B model with the base Qwen3-4B Instruct model across 20 general-domain and biomedical benchmarks drawn from established suites.

Because fine-tuning can introduce risks such as catastrophic forgetting (loss of previously acquired abilities) or mode/label collapse (overly narrow or repetitive outputs), our goal was not exhaustive benchmarking but sensitive detection of any degradation in general or biomedical knowledge. We therefore selected a lightweight yet diverse suite of tasks, combining the HELM-Lite suite benchmarks^30^ with selected standard evaluation tasks (e.g., reading comprehension, factual question answering, and logical reasoning), as well as all publicly available benchmarks from the MedHELM biomedical suite.^5^

Table 4 summarizes the evaluated benchmarks and their targeted abilities^‡^ We interpreted cases in which the fine-tuned model did not exceed the baseline but remained within 7 points of the baseline’s performance as evidence that the trained model retains its original capability on those tasks.

Taken together with the diagnosis prediction and cardiac arrest detection experiments, this benchmark evaluation protocol is intended to test whether a clinically specialized model can preserve broad general-language understanding and reasoning abilities, rather than becoming narrowly tuned to medical content alone. This is particularly important because key sections of the clinical record—such as the history of present illness and goals-of-care discussions—rely heavily on everyday, non-technical language to convey patients’ experiences, preferences, and context, all of which are essential for accurate clinical assessment.

## 4. Results

We evaluated the impact of supervised continued pretraining on clinical notes along three complementary axes. First, we examined the clinical quality of model-generated MDMs through expert review. Second, we quantified knowledge transfer to downstream EHR-based tasks that are closely related to, or more distant from, the MDM generation objective (assigned-diagnosis prediction and in-hospital cardiac arrest detection). Third, we assessed whether clinical specialization altered the model’s broader capabilities using a suite of biomedical and general-domain benchmarks. We detail our results next:

### 4.1. Clinical review of generated MDMs

In Table 1, we report the mean score, obtained by averaging the five quality dimensions across the two reviewers, for the MDMs corresponding to the 25 patient presentations evaluated by our experts. Notably, the scores for the gold standard (usual-care) MDMs reflect the constraints of clinical practice: these notes are generally well written—accurate and internally consistent with the available data—but, due to time pressure, they tend to be brief and sometimes incomplete, particularly regarding the full set of disease processes described in the MDM as differential diagnoses.

**Table 1:** Manual dual-physician expert review of gold-standard and model-generated MDMs for 25 distinct patient presentations. Overall mean scores were computed by averaging the five quality dimensions across two independent reviewers. *(Acc: Accuracy, Compl: Completeness, Use:Usefulness, Cons: Internal Consistency, Compr: Comprehensibility; Scores range from 1 (poor) to 5 (excellent)*.

|  | Acc | Compl | Use | Cons | Compr |
| --- | --- | --- | --- | --- | --- |
| Gold standard Physician | 4.54 | 4.14 | 4.36 | 4.76 | 4.34 |
| Qwen 3 4B off-the-shelf instruct | 4.18 | 4.48 | 3.78 | 4.26 | 4.24 |
| Qwen 3 4B SFT on MDMs | 4.22 | 4.10 | 4.24 | 4.38 | 4.42 |

Our physician reviewers observed that the trained model (Qwen3-4B SFT on MDMs + SFT on IHCA, line 4) showed substantial performance improvement over the baseline model (Qwen 3 4B off-the-shelf instruct, line 3). It closely mimicked the style of the gold standard MDMs, achieving a high usefulness score (4.24) and the highest comprehensibility score (4.42) compared with both the baseline untrained model and the gold standard. However, by mimicking physicians, it has also learned to write short and incomplete MDMs that lack thorough discussions of the differential diagnosis, as shown by the smaller score of completeness (4.10). It also inserts some unfounded claims, resulting in lower internal consistency than that of physicians. Overall, our physicians frequently preferred the MDMs generated by the trained model even over those in the gold standard as it learned to produce professional and more comprehensible MDMs and produced them consistently, and thus scored better than those written by the physicians themselves. While the base model achieved the highest completeness score, it tended to over-explain its reasoning at the expense of focus, often including extraneous content that an attending physician would not include.

### 4.2. Quantitative improvement

We report in Table 2 and Table 3 the performance of our model and the baseline systems on the diagnosis prediction and in-hospital cardiac arrest detection tasks, respectively. For diagnosis prediction, direct baseline (line 1) was substantially outperformed by our model (line 2) — the same architecture but trained to generate MDMs — achieving gains of 3.9 and 16 points in the micro-strict and micro-overlapping F1-scores, respectively. Our model also surpassed larger models (lines 3 and 4), which exhibit poor performance on this task with F1-scores below 5%. For cardiac arrest detection, our model (line 2) performs modestly when applied zero-shot to this new task, obtaining only 10% F1. However, after supervised fine-tuning on the task-specific training set (line 4), the model outperforms its direct competitor (line 3) by 13 F1 points, and further exceeds the leading system (line 5) by 2 F1 points, despite being 101× smaller.

**Table 2:** Comparative performance of the evaluated systems on the assigned diagnosis prediction task. *msPrec*, *msRec*, and *msF1* refer to the micro-strict Precision, Recall, and F1-score, respectively; *moPrec*, *moRec*, and *moF1* refer to the corresponding micro-overlapping metrics. (Systems 1, 3, and 4 are the off-the-shelf instruct models.)

|  | msPrec. | msRec. | msF1 | moPrec. | moRec. | moF1 |
| --- | --- | --- | --- | --- | --- | --- |
| 1. Qwen 3 4B | 0.0165 | 0.0497 | 0.0247 | 0.0484 | 0.1462 | 0.0728 |
| 2. Qwen 3 4B SFT on MDMs | **0.0613** | 0.0658 | **0.0635** | **0.2248** | **0.2412** | **0.2327** |
| 3. Qwen 3 32B | 0.016 | **0.1098** | 0.0279 | 0.0201 | 0.1382 | 0.0351 |
| 4. Llama 3.1 405B | 0.0198 | 0.1059 | 0.0333 | 0.0205 | 0.1098 | 0.0346 |

**Table 3:**
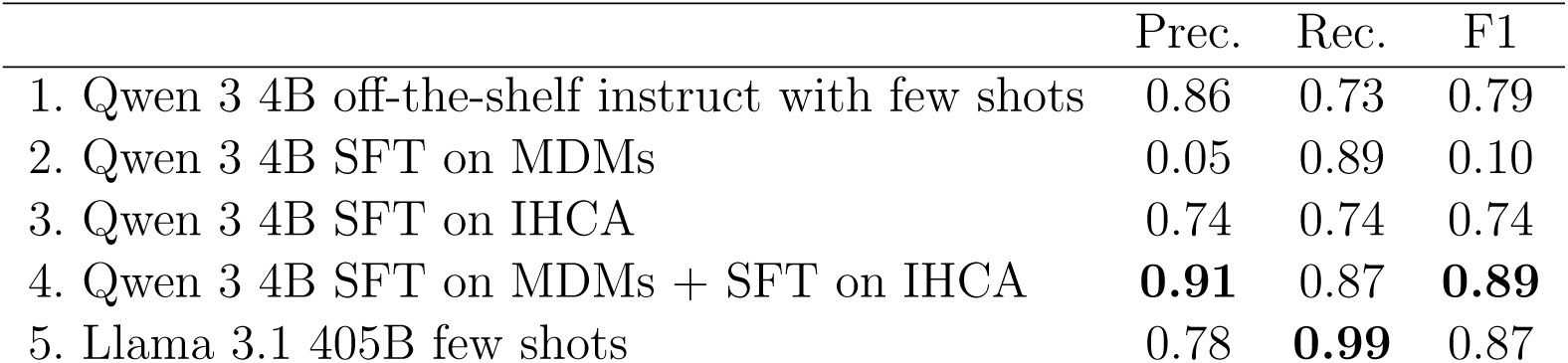
Comparative performance of the evaluated systems on the In-Hospital Cardiac Arrest mentions detection task. Prec., Rec., and F1 refer to the Precision, Recall and F1-score, respectively.

Our results (Table 4) show that the baseline model outperforms the trained model on 10 general-domain and 6 biomedical-domain tasks. Notably, the largest performance drops were observed on multi-step reasoning benchmarks such as GSM8K and MedQA, foreshadowing the reasoning and chain-of-thought limitations we analyze in the Discussion. Nevertheless, the trained model maintains a positive retention rate across both domains, and even surpasses the baseline on select tasks, suggesting that while some performance degradation occurs, the model mainly retains its general knowledge and instruction-following capabilities.

**Table 4:** Impact of fine-tuning on the baseline model’s general and biomedical knowledge performance. The “Performance Retention Rate” reports the proportion of benchmarks where the fine-tuned model outperformed the baseline (bold) or fell within an acceptable difference of 7% (italic). (* evaluation conducted with LightEval using 3-shot learning).

| Benchmark | Qwen 3 4B | Qwen 3 4B | Task Category |
| --- | --- | --- | --- |
|  | off-the-shelf instruct | SFT on MDMs |  |
| Benchmark | Score | Score | Task Category |
| NarrativeQA | **0.723** | 0.606 | Reading-comprehension [QA] |
| NaturalQuestions<br>(Closedbook) | **0.186** | 0.086 | General-knowledge [QA] |
| OpenbookQA | **0.794** | *0.766* | Reading-comprehension [MCQA] |
| MMLU*<br>(49 subjects) | **0.745** | 0.647 | General-knowledge [MCQA] |
| gsm8k* | **0.921** | 0.463 | Math/logic [QA] |
| LegalBench | **0.627** | 0.348 | Legal [MCQA] |
| WMT14 | **0.156** | *0.109* | Translation |
| Arc* | 0.452 | **0.513** | Commonsense reasoning [MCQA] |
| hellaswag* | 0.686 | **0.735** | Commonsense reasoning [MCQA] |
| winogrande* | **0.606** | *0.571* | Commonsense reasoning [MCQA] |
| boolq* | **0.852** | *0.840* | Reading-comprehension [MCQA] |
| truthfulqa (mc1)* | **0.359** | *0.322* | Truthfulness [MCQA] |
| **General domain - Performance Retention Rate: 7/12** |  |  |  |
| MedCalc | **0.082** | *0.051* | Computational Reasoning |
| Medec | **0.524** | *0.521* | Error correction |
| Medbullets | 0.201 | **0.328** | Clinical-comprehension [MCQA] |
| MedQA | **0.555** | 0.408 | Clinical-comprehension [MCQA] |
| MedMCQA* | **0.559** | 0.465 | Clinical-knowledge [MCQA] |
| PubMedQA | 0.548 | **0.562** | Biomedical-comprehension [MCQA] |
| RaceBias | 0.515 | **0.677** | Safeness [MCQA] |
| MedHallu | **0.795** | 0.572 | Truthfulness [MCQA] |
| MMLU*<br>(8 subjects) | **0.750** | 0.658 | Clinical-knowledge [MCQA] |
| **Biomedical domain - Performance Retention Rate: 5/9** |  |  |  |

## 5. Discussion

This study examined whether supervised continued pretraining on real clinical notes can endow an open-source instruction-tuned LLM with richer medical knowledge while preserving its broader language capabilities. Fine-tuning on ED MDM generation altered the model’s outputs in ways that clinicians generally preferred over the base model, while also revealing limitations in completeness and factual grounding that constrain immediate clinical use. Quantitatively, the fine-tuned model transferred its acquired knowledge to downstream clinical tasks, improving assigned-diagnosis prediction and enabling more efficient adaptation to in-hospital cardiac arrest detection compared with both its base counterpart and larger, clinically untrained models. Evaluations on general-purpose and biomedical benchmarks further suggest that, under a conservative fine-tuning regime, clinical specialization need not cause wholesale degradation of non-medical language understanding, an essential property for models that must interpret both technical and narrative aspects of the clinical record. We reflect on each of these aspects next.

### Qualitative evaluation

During a follow-up debriefing, the physicians in our team confirmed that our objective was clearly met: the fine-tuned model outperforms the baseline model on four of the five PDQI-9 dimensions and closely reproduces the style of MDMs documented in our institution’s EHR, including the brevity, implicit reasoning, and communication style that is typical of real-world clinical notes. By contrast, the baseline model produced longer and more explicit reasoning chains—a behavior our physicians valued for comprehensiveness, even though such verbosity is unrealistic and more representative of how a trainee might enumerate all the possible disease processes, rather than selecting the most likely or most consequential ones that experienced clinicians would be expected to document. At the same time, reviewers noted that the fine-tuned model sometimes omitted elements of the differential diagnosis and occasionally introduced weakly supported statements, underscoring that improved style and perceived usefulness do not, by themselves, guarantee clinically adequate or fully reliable documentation.

Further improving the quality of automatically generated MDMs to align with human performance and preferences—particularly by enhancing clinical explanations or making reasoning more explicit—would require methods beyond from our proxy-task training. Achieving this would likely necessitate dedicated alignment procedures, such as reinforcement learning with human feedback, which underpins recent advances in general-purpose chatbots like Chat-GPT.^32^ Such training would also demand new forms of supervision, as clinical notes routinely contain incomplete, implicit, or undocumented aspects of physicians’ reasoning.

One promising research direction involves making this reasoning explicit using formal logic to represent the clinical thought process in a structured, step-wise format. Early prototypes of such approaches are emerging—for example, the work of Freitas et al.**^?^** and our newly developed datasets that explicitly include clinical reasoning steps**^?^**—and suggest a viable path toward future models capable of generating clinically faithful and fully articulated medical decision-making narratives.

### Quantitative evaluation

In the quantitative evaluation, we showed that the model not only acquired medical knowledge during training but also successfully transferred it to two clinical tasks: (1) assigned diagnosis prediction, which is closely related to its training objective, and (2) in-hospital cardiac arrest detection, which represents a more distant task. In both cases, our model outperformed all competing baselines. On the diagnosis prediction task, without any additional training, our model achieved a 16-point F1-score improvement and even surpassed larger models that, in theory, should have performed better due to their size. On the cardiac arrest detection task, while the model struggled to generalize to a completely new task without examples, supervised fine-tuning on that specific dataset enabled it to outperform all competing systems. By comparing our model — first trained on MDM generation and then fine-tuned for cardiac arrest detection — with an identical model trained only on the latter task, we quantified a 15-point F1-score improvement attributable directly to the prior MDM training.

Supervised fine-tuning, while beneficial, also introduced several unintended behaviors. After training, we observed mild mode collapse in multiple tasks. For MDM generation, the model repeatedly produced sentences denying the presence of common findings (*e.g.* repeating “The pt has no appendicitis. The pt has no appendicitis. The pt […]”); for diagnosis prediction, it occasionally output the same ICD-10 codes multiple times. We mitigated these issues during inference by penalizing repetitions of the same sequence of tokens (setting the repetition penalty to 1.2 in the OpenAI API). In the in-hospital cardiac arrest detection task, the model exhibited label collapse, predicting a cardiac arrest in the emergency department for nearly all notes — a pattern likely attributable to the prior MDM-generation training. Although subsequent supervised fine-tuning on the cardiac arrest dataset corrected the problem, this is not an ideal solution, as it requires additional labeled data and substantial computation, underscoring the need for more principled strategies to prevent collapse during clinical specialization.

### Benchmark evaluation

The standard practices recommended to mitigate catastrophic forgetting — namely, incorporating general-domain examples and reducing the learning rate — proved effective, as our model mostly retains its ability to answer a diverse range of questions and remains within a modest performance margin of the base model on many general and biomedical benchmarks. At the same time, manual inspection of outputs on tasks where the base model outperforms the fine-tuned models revealed patterns of label collapse, echoing the behavior we previously observed on the in-hospital cardiac arrest task and indicating that clinical specialization can increase the prior probability of a small set of responses even in non-clinical settings.

Our manual examination of the trained model’s outputs also reveals a reasoning collapse that likely explains its larger performance drop on benchmarks requiring multi-step reasoning, such as *GSM8K* and *MedQA*. Qwen3-4B was originally trained using reinforcement learning to generate an explicit chain of thought, implemented through a <THINK> tag in which the model elaborates its reasoning before closing the tag and generating its response. However, our supervised fine-tuning data — both the medical training set and the 10% general-domain examples included to mitigate catastrophic forgetting — contained no chain-of-thought annotations preceding the target outputs. As a consequence, the model learned to answer directly, and while it still generates the <THINK> tags, their content is either empty or substantially shorter than those produced by the base model. Deprived of meaningful reasoning traces, the trained model fails to leverage its chain-of-thought capability, resulting in degraded performance relative to the baseline on reasoning-intensive tasks. This behavior has direct clinical implications: a model that cannot reliably articulate intermediate steps may still produce fluent but poorly justified conclusions, which are difficult for clinicians to scrutinize.

A natural remedy would be to use a large teacher model to generate chain-of-thought annotations for the gold standard answers of the general-domain training sets, thereby reintroducing reasoning supervision without requiring additional human annotation.

## 6. Limitations and future work

Although we successfully injected medical knowledge into the LLM, its overall performance on MDM generation and diagnosis assignment remained limited, which led us to restrict qualitative evaluation to a small sample, as broader review would not have justified the additional burden on highly specialized clinicians. Several factors contribute to these suboptimal results; however, most of them can potentially be addressed in future work.

Qwen3-4B is a relatively small model, and its limited performance compared to larger LLMs is well documented in the literature.^2,33^ We intentionally selected this model size to validate the feasibility of our approach under realistic resource constraints; however, the modest capacity of a 4B-parameter system inevitably constrains downstream performance. While training trillion-parameter models is beyond our current computational budget, mid-scale models (tens to hundreds of billions of parameters) remain achievable. We are currently training a Qwen 3 14B model, and our physician evaluator has already reviewed MDMs generated by an intermediate checkpoint trained on 400K notes, confirming a noticeable improvement in the richness and clinical relevance of the generated content for 10 patient presentations included in our qualitative analysis.

Improving performance at larger scales, will also require a stronger training strategy rather than relying on supervised full fine-tuning. A key priority is expanding the training corpus: for MDM generation, we are developing a segmenter based on our trained model to automatically extract MDM and assigned-diagnosis sections from notes missed by our regular-expression pipeline, which we estimate will add approximately 4.5 million additional notes to our training set. We also plan to incorporate discharge summaries from the MIMIC-IV dataset^13^ to reduce institution-specific bias. Beyond data expansion, we plan to explore training approaches that go beyond pattern imitation: supervised fine-tuning encourages the model to mimic how clinicians currently write, which can limit its ability to generate more explicit or complete reasoning. To address this, we will investigate reinforcement learning and engage additional physicians to provide structured human feedback on how notes and task-specific outputs should ideally be written. Such approaches may help the model align more closely with expert expectations while supporting more robust generalization across clinical tasks.

A significant limitation of our work is the restricted amount of patient information provided to the LLM during training. First, our de-identification process removed all numerical values, including vital signs and laboratory results, which clinicians routinely scrutinize to identify abnormalities. These values are central to constructing differential diagnoses—the core reasoning process underlying MDM writing—and more broadly to medical decision-making. We are working with our institution to incorporate LLMs into the de-identification pipeline so that only true personal health information is removed, allowing us to retain all clinically relevant numerical information. Second, when evaluating a new ED patient, physicians routinely review prior notes in the EHR to construct a more complete medical history,^34^ whereas our model has access only to the current ED note and not to the patient’s longitudinal record, placing it at a disadvantage relative to the clinicians who authored the original MDMs.

Finally, our current training pipeline does not explicitly supervise chain-of-thought (CoT) reasoning, and we observed a collapse of the base model’ s CoT behavior on multi-step reasoning tasks. In future work, CoT-augmented fine-tuning will need to be balanced against computational cost and the risk of over-constraining outputs, and complemented by targeted strategies to reduce non-reasoning label collapse (for example, through regularization or lightweight task-specific calibration), so that clinical specialization improves medical competence without undermining the diversity or transparency of model behavior.

## 7. Conclusion

Privacy constraints are currently limiting the amount of real clinical text available for training open-source LLMs, leaving these models with insufficient medical knowledge for reliable clinical use. Using 500K de-identified emergency department physician notes from Cedars-Sinai Medical Center, this study shows that fully supervised fine-tuning of a compact open-source instruction model on a straightforward medical decision-making prediction task can substantially narrow this gap, producing MDMs that clinicians often prefer stylistically. We also showed that our fine-tuned model had better performance on assigned-diagnosis prediction and in-hospital cardiac arrest detection relative to both its base counterpart and larger clinically untrained models. Through qualitative and quantitative evaluations across multiple clinical and general-domain tasks, we show that this approach enables the model to acquire and transfer clinically relevant knowledge, improving performance on unseen biomedical tasks while preserving its general conversational abilities. At the same time, the emergence of mode and label collapse, along with degradation of chain-of-thought behavior on reasoning-intensive benchmarks, underscores that näıve supervised continued pretraining is insufficient to guarantee safe or fully reliable clinical deployment. Overall, our benchmark results indicate that, under a conservative training regime, clinical specialization can be achieved without wholesale loss of general-domain capabilities, suggesting a viable path toward EHR-informed, institution-scale models that support real-world decision-making while retaining the breadth needed to interpret the heterogeneous narratives that characterize routine care.

## Data Availability

As the data were de-identified using an automated process, residual identification risks cannot be fully excluded. Therefore, the dataset and models will not be publicly released to protect patient privacy.

## Acknowledgments

The research reported in this publication was supported by a Centers for Disease Control and Prevention (CDC) Cooperative Agreement Funding Opportunity Announcement (FOA) U54 CK000610, Epicenters for the Prevention of Healthcare-Associated Infections. Dr. Berdahl receives salary support from the Agency for Healthcare Research and Quality under K08HS029534, and he is also supported by the Emergency Medicine Foundation award on Diagnostic Excellence.

∗ The 10% is calculated based on the total number of tokens in the MDM sections of the notes in our training set, excluding the prompt tokens.

† We also accepted ICD-9 codes when they were correctly predicted.

‡ Some HELM-Lite and MedHELM tasks could not be run with their recommended instructions. When this occurred, we attempted to run them through the LightEval benchmarks suite^31^ with 3-shot prompting. In cases where the post-processing steps provided by either framework failed to recover coherent answers from model outputs, we resorted to a custom evaluation interface, employing guided decoding to ensure consistent and accurate parsing. Only tasks that were successfully evaluated are reported in Table 4. For each benchmark, we adopted the default scoring metrics defined by the corresponding suite (e.g., accuracy or exact match for question answering, multiple-choice accuracy for knowledge tasks).^5,29^

